# A pharmacoepidemiological study of myocarditis and pericarditis following mRNA COVID-19 vaccination in Europe

**DOI:** 10.1101/2022.05.27.22275706

**Authors:** Joana Tome, Logan T. Cowan, Isaac Chun-Hai Fung

**Author notes:** Author to whom correspondence should be addressed. Isaac Chun-Hai Fung, PhD, Department of Biostatistics, Epidemiology and Environmental Health Sciences, Jiann-Ping Hsu College of Public Health, Georgia Southern University, PO Box 7989, Statesboro, GA 30460, USA. Disclaimer: The views expressed in the submitted manuscript are ours and not an official position of any institution or funder.

## Abstract

**Purpose:** To assess myocarditis and pericarditis reporting rate as adverse drug reactions (ADRs) for the messenger ribonucleic acid (mRNA) coronavirus disease 2019 (COVID-19) vaccines authorized in Europe.

**Methods:** Data on myocarditis and pericarditis related to mRNA COVID-19 vaccines (period: January 1, 2021 - February 11, 2022) were collected from the EudraVigilance database and combined with the European Centre for Disease Prevention and Control’s (ECDC) vaccination tracker database. The reporting rate was expressed as 1 million individual vaccinated-days with a corresponding 95% confidence interval (CI), and an observed-to-expected (OE) analysis was performed to check if there was an excess risk for myocarditis or pericarditis following mRNA COVID-19 vaccination.

**Results:** The reporting rate of myocarditis per 1 million individual vaccinated-days in the study period was 17.27 (95% CI, 16.34-18.26) for the CX-024414 vaccine and 8.44 (95% CI, 8.18-8.70) for TOZINAMERAN vaccine. The reporting rate for pericarditis per 1 million individual vaccinated-days in the study period was 9.76 (95% CI, 9.06-10.51) for the CX-024414 vaccine and 5.79 (95% CI, 5.56-6.01) for TOZINAMERAN vaccine. The OE analysis showed that both vaccines produced a myocarditis standardized morbidity ratio (SMR) greater than 1, with the CX-024414 vaccine having a greater SMR than TOZINAMERAN. Regarding TOZINAMERAN, SMR for pericarditis was greater than 1 when considering the lowest background incidence, but smaller than 1 when considering the highest background incidence.

**Conclusions:** Our results suggest an excess risk of myocarditis following the first dose of mRNA COVID-19 vaccine, but the relationship between pericarditis and mRNA COVID-19 vaccine remains unclear.

## INTRODUCTION

Myocarditis (inflammation of the heart muscle) and pericarditis (inflammation of the heart’s outer lining) following immunizations such as smallpox, influenza, hepatitis B, or other vaccines have previously been reported as a rare side effect.^1-3^ Depending on the source, the incidence of myocarditis and pericarditis in the European Economic Area (EEA) has ranged from 1 to 10 in 100,000 people per year.^4^ European Medicines Agency (EMA)’s safety committee, the Pharmacovigilance Risk Assessment Committee (PRAC), started reviewing cases of myocarditis and pericarditis following Pfizer-BioNTech (TOZINAMERAN) and Moderna (CX-024414) COVID-19 vaccination since April 2021. Increased rates of cases of myocarditis and pericarditis following mRNA COVID-19 vaccination were also noticed in the United States^5^ and Canada.^6^

TOZINAMERAN and CX-024414, the two mRNA vaccines being used in the European Union (EU)/EEA, contain an mRNA molecule with instructions for producing the spike protein from severe acute respiratory syndrome coronavirus 2 (SARS-CoV-2) that stimulates the body’s immune response.^4^ There are different biological mechanisms that can explain the possible association between mRNA COVID-19 vaccination and the occurrence of myocarditis and pericarditis. Based on preliminary evidence from adult trials with mRNA vaccinations, it is rare but possible that antibody responses to mRNA vaccines can become very high, causing cardiac inflammation.^7^ One other mechanism is the activation of anti-idiotype cross-reactive antibody-mediated cytokine expression in the myocardium, as well as abnormal apoptosis, which can result in myocardial and pericardial inflammation.^8^ The mRNA vaccines may possibly trigger a non-specific innate inflammatory response or a molecular mimicry mechanism between the viral spike protein and an unknown cardiac protein.^9^ The last explanation is that vaccinations’ potent immunogenic RNA can have a bystander or adjuvant consequence on the heart.^10^

Although the occurrence of myocarditis and pericarditis following SARS-CoV-2 infection seems to be more frequent and serious,^11-13^ assessing the risk of myocarditis and pericarditis after COVID-19 mRNA immunization has its own importance.

Previous studies indicate that the incidence of myocarditis and pericarditis may increase following COVID-19 mRNA immunization;^14-18^ therefore, it is critical to determine the magnitude of the association between myocarditis or pericarditis occurrence and mRNA COVID-19 vaccination in order to protect the most vulnerable population and to aid in the differentiation of the COVID-19 vaccination’s risks and benefits. Moreover, many clinical trials were unable to identify rare adverse events of COVID-19 vaccines because they were underpowered. Their detection is critical for risk-benefit analyses and informing post-vaccination clinical practice. Therefore, detecting such adverse events through pharmacovigilance post-authorization has become a global scientific priority.

To enhance risk communication that accompanies existing COVID-19 vaccination campaigns, this study aims to quantify the elevated risk, if any, of myocarditis and pericarditis 28 days following Tozinameran and CX-024414 vaccination among Europeans using publicly available data from the EU/EEA in an exploratory way. This study used an OE analysis, which is a part of the quantitative pharmacovigilance toolkit for vaccines, to calculate the standardized morbidity ratio (SMR).

## MATERIAL AND METHODS

We hypothesize that myocarditis and pericarditis SMRs greater than 1 will result from both the mRNA COVID-19 vaccines. A combination of two databases was used in this pharmacoepidemiological study. The participants were European people from the EU/EEA countries who were eligible for the mRNA COVID-19 vaccines that were available in Europe during the 2021-2022 periods. All the variables included in this analysis are named/defined as classified in their respective databases.

ECDC has a vaccination tracker database^19^ that gathers reports from the EU/EEA countries. These countries submit data to the ECDC through The European Surveillance System (TESSy) twice a week (Tuesdays and Fridays). The database contains aggregated data on the number of vaccine doses distributed by manufacturers to the EU/EEA countries, the number of first, second, additional, and unspecified doses administered to different age groups and in specific target groups, such as healthcare workers and in residents in long-term care facilities. The database includes data on the COVID-19 vaccination rollout, where each row contains the corresponding data for a certain week and country. The database is publicly available and was downloaded on February 16, 2022. From this database, we extracted the number of individuals from the EU/EEA countries who received their first dose of CX-024414 and TOZINAMERAN vaccines that also represents our exposure variable. Technically, being vaccinated with the first dose of the CX-024414 or TOZINAMERAN vaccines (including a time window from the date when the mRNA COVID-19 vaccines became eligible for vaccination in Europe till February 16, 2022) is considered as our exposure variable. In the analysis, it was used as the sum of the individuals from the EU/EEA countries who received their first dose of CX-024414 and TOZINAMERAN vaccines during the time window period, which also represents our study population size.

EudraVigilance^20^ is a public available pharmacovigilance database of suspected ADR reports. Data was downloaded from EudraVigilance on February 16, 2022. The database contains cardiac problems related to COVID-19 vaccination using mRNA vaccines (CX-024414 and TOZINAMERAN) reported from January 1, 2021, to February 11, 2022. Among all cardiac problems, our outcomes of interest are myocarditis and pericarditis. The database contains the EU local number (identification number), type of report, the date when the report was received, the source of the report, age group, sex, the vaccine type, the adverse event, the duration, outcome, and seriousness of the adverse event, the duration, dose, and the route of administration of the vaccine, the type of concomitant, and the duration, dose, and the route of administration of any concomitant received by the patient. In this analysis, we used the number of myocarditis and pericarditis ADRs, the age group, sex, and the vaccine as the other needed variables. The ADR outcomes were treated as the sum of the number of myocarditis or pericarditis reported in the study period. Age group categories were classified as 5-11, 12-17, 18-64, 65-85 years old, more than 85 years old, and not specified. Sex was classified as female, male, and not specified. The vaccine variable was represented by the CX-024414 and TOZINAMERAN vaccines. No missing data were observed from both the databases.

Myocarditis and pericarditis reporting rates during the study period (January 1, 2021, to February 11, 2022) were estimated using the formulae: ^21^

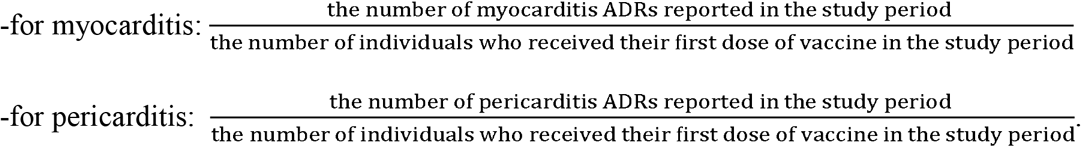

The OE analysis was performed within 28 days post the first vaccine dose, considering the estimated lowest and highest myocarditis and pericarditis background incidence rates reported by the EMA.^4^ The background incidence rate represents the number of new cases naturally occurring in the EEA, expressed as person-time, and was estimated for the EEA population before it was exposed to any of the mRNA COVID-19 vaccines. The person-time at risk was calculated as the number of first doses × (28/365.2425) × (1/100,000).^21^ The OE ratio was expressed as an SMR with a 95% CI.^21^ The analysis was performed using the ‘epiR’ package in R version 4.0.3 (R Core Team, R Foundation for Statistical Computing, Vienna, Austria).

### Patient and public involvement

Only publicly available data are used in this study. This study was determined by Georgia Southern University’s Institution Review Board (IRB) (H20364) to be exempt from full review under the G8 exemption category (Non-human subjects determination): This project does not involve obtaining information about living individuals or does not have direct interaction or intervention with individuals or their personal data so is not defined as human subjects research under human subjects regulations. As well, no patient or public was involved in the study design or the data collection, analysis, or interpretation procedures.

## RESULTS

In the study period, there were 73,466,253 people in the EU who received the first dose of the CX-024414 vaccine and 484,402,251 who received their first dose of the TOZINAMERAN vaccine. In the same period, there were 9,078 and 38,297 individuals in EU/EEA countries reported with adverse cardiac problems after the administration of the CX-024414 vaccine and the TOZINAMERAN vaccine, respectively. Of these reports, there were 1,269 and 4,087 myocarditis events reported for CX-024414, and for TOZINAMERAN respectively (**Table 1**), and 717 and 2,803 pericarditis events for CX-024414 and TOZINAMERAN, respectively (**Table 2**). The reporting rate of myocarditis per 1 million individual vaccinated-days in the study period was 17.27 (95% CI, 16.34-18.26) for the CX-024414 vaccine and 8.44 (95% CI, 8.18-8.70) for TOZINAMERAN vaccine. The reporting rate for pericarditis per 1 million individual vaccinated-days in the study period was 9.76 (95% CI, 9.06-10.51) for the CX-024414 vaccine and 5.79 (95% CI, 5.56-6.01) for TOZINAMERAN vaccine.

**Table 1.**
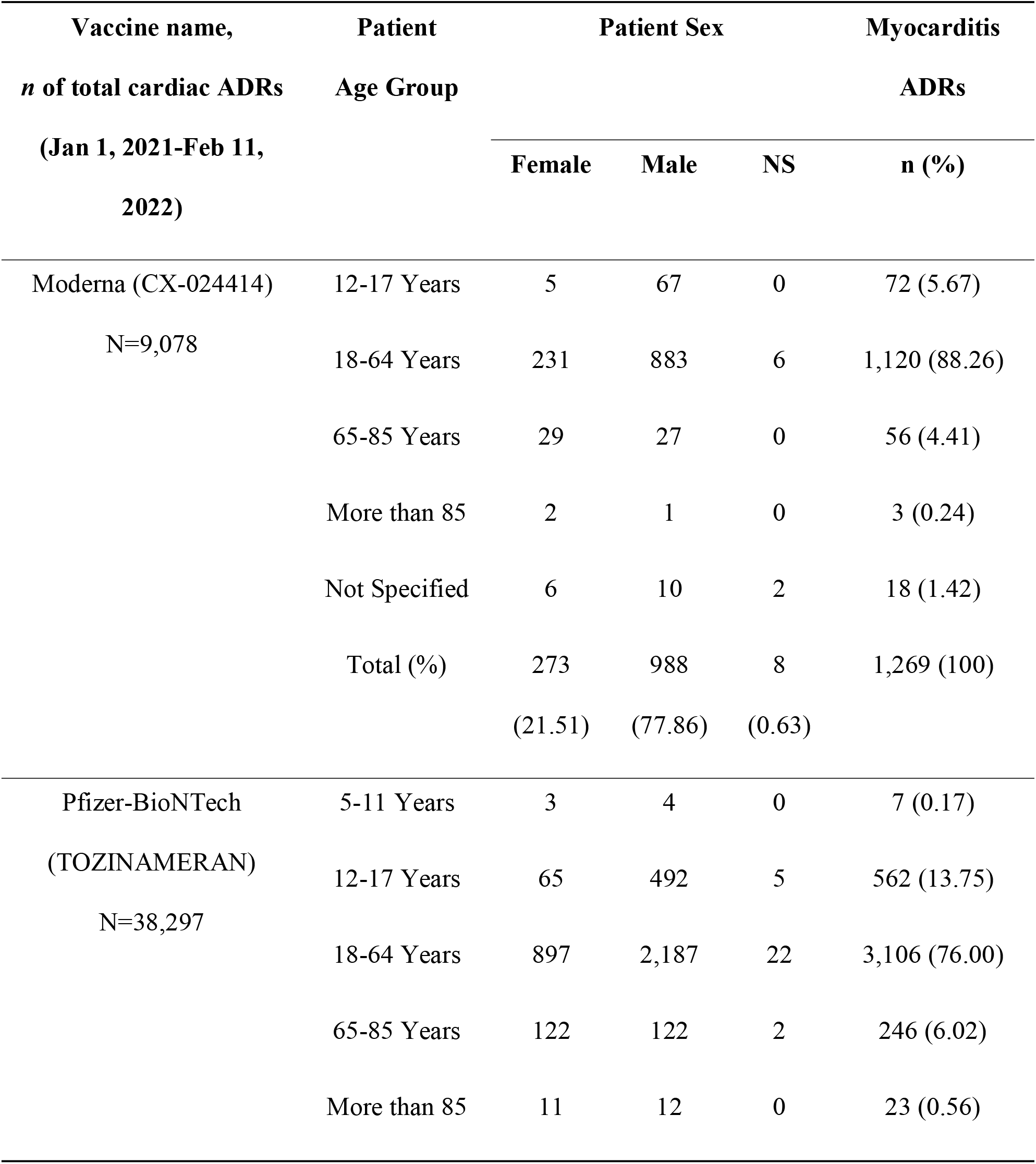
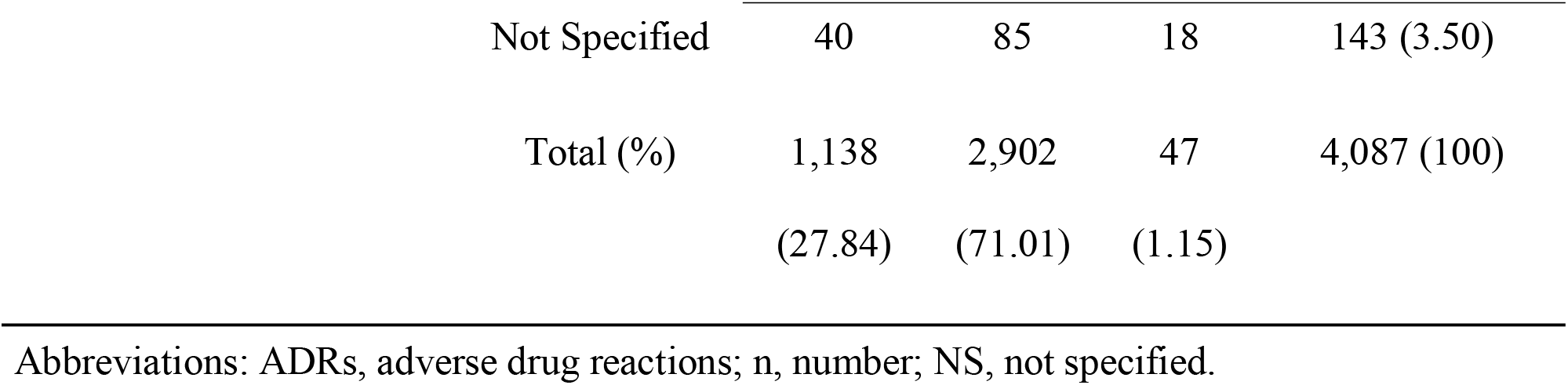
All myocarditis ADRs stratified by age group and sex.

| Vaccine name,<br>*n* of total cardiac ADRs<br>(Jan 1, 2021-Feb 11,<br>2022) | Patient<br>Age Group | Patient Sex |  |  | Myocarditis<br>ADRs |
| --- | --- | --- | --- | --- | --- |
|  |  | Female | Male | NS | n (%) |
| Moderna (CX-024414)<br>N=9,078 | 12-17 Years | 5 | 67 | 0 | 72 (5.67) |
|  | 18-64 Years | 231 | 883 | 6 | 1,120 (88.26) |
|  | 65-85 Years | 29 | 27 | 0 | 56 (4.41) |
|  | More than 85 | 2 | 1 | 0 | 3 (0.24) |
|  | Not Specified | 6 | 10 | 2 | 18 (1.42) |
|  | Total (%) | 273<br>(21.51) | 988<br>(77.86) | 8<br>(0.63) | 1,269 (100) |
| Pfizer-BioNTech<br>(TOZINAMERAN)<br>N=38,297 | 5-11 Years | 3 | 4 | 0 | 7 (0.17) |
|  | 12-17 Years | 65 | 492 | 5 | 562 (13.75) |
|  | 18-64 Years | 897 | 2,187 | 22 | 3,106 (76.00) |
|  | 65-85 Years | 122 | 122 | 2 | 246 (6.02) |
|  | More than 85 | 11 | 12 | 0 | 23 (0.56) |

| **Vaccine name,** | **Patient** | **Patient Sex** |  |  | **Myocarditis** |
| --- | --- | --- | --- | --- | --- |
| ***n* of total cardiac ADRs** | **Age Group** |  |  |  | **ADRs** |
| **(Jan 1, 2021-Feb 11, 2022)** |  | **Female** | **Male** | **NS** | **n (%)** |
|  | Not Specified | 40 | 85 | 18 | 143 (3.50) |
|  | Total (%) | 1,138<br>(27.84) | 2,902<br>(71.01) | 47<br>(1.15) | 4,087 (100) |

Abbreviations: ADRs, adverse drug reactions; n, number; NS, not specified.

**Table 2.** All pericarditis ADRs stratified by age group and sex.

| **Vaccine name,** | **Patient** | **Patient Sex** |  |  | **Pericarditis** |
| --- | --- | --- | --- | --- | --- |
| ***n* of total cardiac ADRs** | **Age Group** |  |  |  | **ADRs** |
| **(Jan 1, 2021-Feb 11, 2022)** |  | **Female** | **Male** | **NS** | **n (%)** |
| Moderna (CX-024414) | 12-17 Years | 5 | 7 | 0 | 12 (1.67) |
| N=9,078 | 18-64 Years | 266 | 336 | 4 | 606 (84.52) |
|  | 65-85 Years | 46 | 43 | 0 | 89 (12.41) |
|  | More than 85 | 2 | 0 | 0 | 2 (0.28) |
|  | Not Specified | 4 | 4 | 0 | 8 (1.12) |
|  | Total (%) | 323 | 390 | 4 | 717 |

| Vaccine name,<br>*n* of total cardiac ADRs<br>(Jan 1, 2021-Feb 11,<br>2022) | Patient<br>Age Group | Patient Sex |  |  | Pericarditis |
| --- | --- | --- | --- | --- | --- |
|  |  | Female | Male | NS | ADRs |
|  |  |  |  |  | n (%) |
|  |  | (45.05) | (54.39) | (0.56) |  |
| Pfizer-BioNTech<br>(TOZINAMERAN)<br>N=38,297 | 12-17 Years | 49 | 89 | 0 | 138 (4.92) |
|  | 18-64 Years | 1,157 | 1,068 | 10 | 2,235 (79.74) |
|  | 65-85 Years | 160 | 184 | 0 | 344 (12.27) |
|  | More than 85 | 15 | 25 | 0 | 40 (1.43) |
|  | Not Specified | 24 | 20 | 2 | 46 (1.64) |
|  | Total (%) | 1,405<br>(50.12) | 1,386<br>(49.45) | 12<br>(0.43) | 2,803 |

Abbreviations: ADRs, adverse drug reactions; n, number; NS, not specified.

**Tables 3** and **4** presented the OE analyses for both vaccines using two background incidence rates, namely 1 and 10 per 100,000 persons per year, respectively. For myocarditis, the results showed an SMR >1 for both vaccines, with the CX-024414 vaccine having a greater SMR than that of TOZINAMERAN. For pericarditis, both vaccines had an SMR >1 when considering the lowest background incidence; however, when the highest background incidence was considered, the SMR for TOZINAMERAN was <1 while that for CX-024414 was >1. Still, regarding pericarditis, the CX-024414 vaccine had greater SMRs than those of the TOZINAMERAN vaccine.

**Table 3.** Observed-to-expected analysis for myocarditis.

| **Vaccine name** | **Person-time at risk (100,000 person-years)** | **Background incidence (lowest, highest)** | **Expected Myocarditis** | **Observed Myocarditis** | **SMR (95% CI)** |
| --- | --- | --- | --- | --- | --- |
| Moderna (CX-024414) | 56.32 | 1 | 56.32 | 1,269 | 22.53<br>(21.31-23.81) |
|  |  | 10 | 563.2 |  | 2.253<br>(2.13-2.38) |
| Pfizer-BioNTech (TOZINAMERAN) | 371.35 | 1 | 371.35 | 4,087 | 11.01<br>(10.67-11.35) |
|  |  | 10 | 3,713.5 |  | 1.10<br>(1.07-1.14) |

Abbreviations: SMR, standardized morbidity ratio; CI, confidence interval.

**Table 4.** Observed-to-expected analysis for pericarditis.

| **Vaccine name** | **Person-time at risk (100,000 person-years)** | **Background incidence (lowest, highest)** | **Expected Pericarditis** | **Observed Pericarditis** | **SMR (95% CI)** |
| --- | --- | --- | --- | --- | --- |
| Moderna | 56.32 | 1 | 56.32 | 717 | 12.73 |
| (CX-024414) |  |  |  |  | (11.82-13.70) |
|  |  | 10 | 563.2 |  | 1.27 |
|  |  |  |  |  | (1.18-1.37) |
| Pfizer- | 371.35 | 1 | 371.35 | 2,803 | 7.55 |
| BioNTech |  |  |  |  | (7.27-7.83) |
| (TOZINAME |  | 10 | 3,713.5 |  | 0.75 |
| RAN) |  |  |  |  | (0.73-0.78) |

Abbreviations: SMR, standardized morbidity ratio; CI, confidence interval.

## DISCUSSION

In this study, we used data from EudraVigilance to evaluate the occurrence of myocarditis and pericarditis after mRNA COVID-19 vaccination. The majority of myocarditis and pericarditis’ ADRs were reported among 18-64 years old people. Myocarditis occurred mostly among male cases, while for pericarditis, there were slightly more ADRs among males for CX-024414 and slightly more ADRs among females for the TONZINAMERAN vaccine. However, these results should be interpreted cautiously because data on the vaccination rate stratified by age and sex are absent in this study. The reporting rate of myocarditis and pericarditis per 1 million individual vaccinated-days in the study period was greater for CX-024414 compared to TOZINAMERAN. The SMR for myocarditis was greater than 1 for both vaccines (showing excess risk), with the CX-024414 vaccine having a greater SMR than TOZINAMERAN. For pericarditis, the results showed an SMR > 1 when considering the lowest background incidence; however, when the highest background incidence was considered, the TOZINAMERAN vaccine showed a smaller than 1 SMR. In either case, the pericarditis SMRs for CX-024414 were greater than that for TOZINAMERAN.

In Germany, surveillance data from the Paul Ehrlich Institute reported a rate for myocarditis in men 18-29 years old of 11.71 cases per 100,000 COVID-19 Moderna vaccine recipients in German data and 7.79 in the Moderna Global Safety Database.^22^ Furthermore, two other large European studies reported the occurrence of myocarditis and pericarditis after COVID-19 mRNA vaccination.^22^ The first one, the Nordic cohort study of 23 million residents of Denmark, Finland, Norway, and Sweden, observed that during the 28-day risk periods following vaccination, 1,092 incident myocarditis cases and 1,154 incident pericarditis cases occurred. Among them, 106 and 123 myocarditis cases occurred following first-and second-dose vaccinations with TOZINAMERAN, respectively, and 15 and 67 following CX-024414, respectively. The second one, the population-based case-control study in France, ^22^ showed a large increase in the myocarditis odds within seven days after vaccination. In this study, the association with the risk of myocarditis appeared particularly pronounced in young men under 30 years of age, particularly after the second dose of the CX-024414 vaccine (odds ratio (OR) 79.8; 95% CI [29.8-213.4]). A Danish population-based cohort study observed that vaccination with the Moderna vaccine was significantly associated with an increased risk of myocarditis among 12-39 years old people, while vaccination with the Pfizer-BioNTech vaccine was significantly associated with an increased risk among women.^23^

Meanwhile, it is noteworthy to mention that the risk of myocarditis among adolescents was found in one study to be lower after a booster compared to the second dose of the Pfizer-BioNTech COVID-19 vaccine. ^24^

While an elevated risk of myocarditis following vaccination with mRNA COVID-19 vaccine was observed, the evidence regarding pericarditis remained equivocal. The population-based case-control study in France observed an increased risk of pericarditis in people under 30 years of age, in particular after the second dose in men (OR 15.0, 95% CI [3.3-68.4]) and after the first dose in women (OR 27.9, 95% CI [2.4-328.0]), ^22^ while the Danish population-based cohort study observed an increased risk of pericarditis among 12-39 years old people. ^23^ On the other hand, Patone et al.^25^ observed that there was no evidence of an increased risk of pericarditis following vaccination, except in 1 to 28 days after the second dose of the Moderna vaccine.

Furthermore, Diaz et al.^18^ reported that pericarditis affected older patients later, after either the first or second dose.

The elevated risk of myocarditis (and perhaps pericarditis) after receiving mRNA COVID-19 vaccine should be evaluated in light of the greater risk of myocarditis or pericarditis following SARS-CoV-2 infection.^25^ Oberweis et al.^26^ suggested that cardiac injury from SARS-CoV-2 can be caused directly (e.g., through angiotensin-converting enzyme 2 binding in the heart) and indirectly through a toxic inflammatory reaction with cytokine storm, with the indirect mechanism being more frequent. Furthermore, the authors hypothesized that some children have died from COVID-19 because they may have been more susceptible to heart damage from the cytokine storm caused by COVID-19 than the standard respiratory distress syndrome seen in adults.

Even though the rare occurrence of myocarditis and pericarditis can be deadly, the majority of myocarditis cases linked to vaccines have been mild and self-limiting,^17^ whereas SARS-CoV-2 infection may carry a serious risk of morbidity and mortality over time, especially for the unvaccinated. Myocardial injury was highly common among hospitalized COVID-19 patients.^27^ Patone et al.^25^ emphasized that compared to the risk from COVID-19 vaccine, SARS-CoV-2 infection was associated to a significant increase in the risk of myocarditis, pericarditis, and cardiac arrhythmia-related hospitalization or death. Therefore, the benefits of SARS-CoV-2 mRNA vaccination that lowers both the risk of infection and the risk of hospitalization should be considered when interpreting the results of our study.

Furthermore, our results should be interpreted in light of the fact that myocarditis and pericarditis have also occurred following immunizations such as smallpox, influenza, hepatitis B, or other vaccines.^1-3^

Our study is subject to certain limitations. First, we cannot directly compare the reporting rate of myocarditis or pericarditis to the incidence rate in the general EU/EEA population because they represent different measures that use different definitions of the time at risk.

Consequently, whether the incidence of myocarditis or pericarditis is higher in vaccinated people than in the general population is beyond the scope of this study. Since we do not know the exact background incidence rate, which may vary substantially among different vaccine group populations, as well as knowing that the data were based on different assumptions,^28^ the findings should be interpreted with caution. Second, we cannot exclude the possibility of ADRs’ registration underestimation. There is a possibility of underreporting myocarditis and pericarditis, which can impose non-differential misclassification. Third, we were unable to calculate myocarditis and pericarditis incidence for each vaccine that is adjusted for demographics and other factors because the information on confounding is absent; except for sex and age group, information on other variables was missing. Fourth, the reporting rates were not standardized for age due to the unavailability of the data; therefore, we were unable to perform a stratified OE analysis. A vaccine’s safety profile may vary depending on the target population (e.g., higher risks in the youngest age groups); therefore, comparing reporting rates for age groups or countries should be avoided because it would introduce biases and inaccuracies.

## CONCLUSION

To conclude, our study analysis supports an excess risk of myocarditis following the first dose of the mRNA COVID-19 vaccine, but the direction of the relationship between pericarditis and mRNA COVID-19 vaccine remains unclear. Future research is needed to calculate age and sex standardized reporting rates, stratified OE analyses, as well as analyses following the second or more doses of mRNA COVID-19 vaccine. The benefits of COVID19 vaccination exceed the risks of adverse events; however, pharmacovigilance methods continue to play a pivotal role in monitoring, identifying, and reducing ADR risk.

## Data Availability

All data produced in the present work are contained in the manuscript.

## ABBREVIATIONS

ADRs: adverse drug reactions
CI: confidence interval
COVID-19: coronavirus disease 2019
ECDC: European Centre for Disease Prevention and Control
EEA: European Economic Area
EMA: European Medicines Agency
EU: European Union
mRNA: messenger ribonucleic acid
OE: observed-to-expected
OR: odds ratio
PRAC: Pharmacovigilance Risk Assessment Committee
RNA: ribonucleic acid
SARS-CoV-2: severe acute respiratory syndrome coronavirus 2
SMR: standardized morbidity ratio

## ACKNOWLEDGEMENTS

None.

## CONFLICT OF INTERESTS

None declared.

## FUNDING

This research did not receive any specific grant from funding agencies in the public, commercial, or not-for-profit sectors.

## CONTRIBUTORSHIP

Joana Tome – Research question generation, literature review, data analysis and interpretation, manuscript preparation.

Logan T. Cowan - Manuscript preparation and review.

Isaac Chun-Hai Fung - Literature review, manuscript preparation, and review.

All authors attest they meet the ICMJE criteria for authorship.

## Notes

Source(s) of support: None

### Competing Interest Statement

The authors have declared no competing interest.

### Funding Statement

This study did not receive any funding.

### Author Declarations

Only publicly available data are used in this study. This study was determined by Georgia Southern University's Institution Review Board (IRB) (H20364) to be exempt from full review under the G8 exemption category (Non-human subjects determination): This project does not involve obtaining information about living individuals or does not have direct interaction or intervention with individuals or their personal data so is not defined as human subjects research under human subjects regulations. ECDC Data on COVID-19 vaccination in the EU/EEA. Available at: https://www.ecdc.europa.eu/en/publications-data/data-covid-19-vaccination-eu-eea. Accessed 2/16/2022. EudraVigilance: European Database of suspected adverse drug reaction. European Medicines Agency. Available at: https://www.adrreports.eu/. Accessed 2/16/2022.

### Summary of Updates

A minor edit was made to the abstract to clarify the meaning of a sentence.

